# Developing a context-relevant psychosocial stimulation intervention to promote cognitive development of children with severe acute malnutrition in Mwanza, Tanzania

**DOI:** 10.1101/2023.04.19.23288798

**Authors:** C. L. Jensen, E. Sanga, H. Kitt, G. PrayGod, H. Kunzi, T. Setebe, S. Filteau, J. Webster, M. Gladstone, M. F. Olsen

## Abstract

More than 250 million children will not meet their developmental potential due to poverty and malnutrition. Psychosocial stimulation (PS) has shown promising effects for improving development in children exposed to severe acute malnutrition (SAM) but programs are rarely implemented. In this study, we used qualitative methods to inform the development of a PS programme to be integrated with SAM treatment in Mwanza, Tanzania. We conducted in-depth interviews with seven caregivers of children recently treated for SAM and nine professionals in early child development. We used thematic content analysis and group feedback sessions and organised our results within the Nurturing Care Framework. Common barriers to stimulate child development included financial and food insecurity, competing time demands, low awareness about importance of responsive caregiving and stimulating environment, poor father involvement, and gender inequality. Caregivers and professionals suggested that community-based support after SAM treatment and counselling on PS would be helpful, e.g. how to create homemade toys and stimulate through involvement in everyday chores. Based on the findings of this study we developed a context-relevant PS programme. Some issues identified were structural highlighting the need for programmes to be linked with broader supportive initiatives.

**Key findings:**

- Financial insecurity, competing time constraints, lack of awareness and poor father involvement are all barriers highlighted by families with children who have severe acute malnutrition (SAM) as negatively impacting their ability to promote nurturing care and child development.
- Despite knowledge of the importance of good health and adequate nutrition as important contributors to child development, caregivers of children with SAM did not always have the resources to support this.
- Caregivers interviewed were less familiar with the importance of responsive caregiving and opportunities for early learning as ways to support children’s development.
- A context-relevant programme to support psychosocial development among young children treated for SAM should include clear linkages with community-based support after discharge, but also approaches to promote involvement of fathers in childcare, and counselling of caregivers on low-cost strategies to improve nutrition and development.
- Other supporting social welfare initiatives in the community are needed to address the broader structural issues identified in this study.

## Introduction

More than 250 million children below 5 years do not reach their full developmental potential, mainly due to poverty and malnutrition (Black *et al*., 2017). Most of these children live in low- and middle-income countries with the highest prevalence found in sub-Saharan Africa (Black *et al*., 2017). Malnutrition is common in these settings and research has demonstrated that both chronic and acute malnutrition impair cognitive and motor development (Abessa *et al*., 2017; Sudfeld *et al*., 2015; van den Heuvel *et al*., 2017). This results in long-term consequences for child development and future life opportunities, as developmental delays track into later childhood and adulthood (Kirolos *et al*., 2022).

Jamaican studies from the 1970s showed that psychosocial stimulation (PS) of malnourished children can improve child development (Grantham-McGregor *et al*., 1980; Grantham-McGregor *et al*., 1987). These studies demonstrated for the first time, that weekly PS improve developmental outcomes in children with severe acute malnutrition (SAM) more than nutritional therapy on its own. More recent studies have confirmed beneficial effects of PS in SAM (Abessa *et al*., 2019; Nahar *et al*., 2012). In addition, programmes in community settings have shown that caregiver support through PS programmes can improve child development and cognition at two years of age (Hamadani *et al*., 2019; Yousafzai *et al*., 2014) – although some, when integrated into routine government programmes, have shown less benefit (Jeong *et al*., 2021).

Based on the early studies in Jamaica, WHO guidelines for inpatient treatment of SAM have included recommendations on PS for the past two decades (Ashworth *et al*., 2003). However, PS activities are rarely implemented in inpatient treatment and still not mentioned in guidelines for community management of SAM (Bahwere *et al*., 2006). More recently, the Nurturing Care Framework (NCF) was created by international agencies (WHO *et al*., 2018) as a framework of actionable strategies to improve early child development (ECD). The NCF describes five components of nurturing care needed for children to reach their full developmental potential: good health, adequate nutrition, responsive caregiving, opportunities for early learning, and security and safety (WHO *et al*., 2018). Programmes have been created to support parents in providing PS, including the Care for Child Development intervention package (WHO & UNICEF, 2012). However, these programs are rarely implemented in management of SAM, despite children being one of the most high-risk groups.

The lack of PS activities in SAM management is likely due to resource-constraints, such as training of staff and supervision (Cavallera *et al*., 2019). We still do not have good evidence as to what factors may facilitate effective implementation of PS programmes for this vulnerable group (Daniel *et al*., 2017; Jeong *et al*., 2021; Mehrin *et al*., 2022). To understand better strategies for implementation and facilitate scale-up, it is crucial to gain the insight of caregivers and professionals who work with SAM and ECD (Mehrin *et al*., 2021; Richter *et al*., 2017). In this study in Mwanza, Tanzania, we aimed to 1) explore barriers and facilitators in promoting PS of children treated for SAM, and 2) to explore perceptions of caregivers and professionals and their suggestions as how best to design a context-specific PS package to be integrated with SAM treatment.

## Methods

### Study design and setting

With a long-term plan of conducting a factorial trial to test the effects of improved nutritional and PS interventions in the management of malnutrition, we conducted the ‘BrightSAM (Brain development, growth and health in children with SAM) trial development study’ between June 2020 – February 2022. The overall aim was to develop, pilot test and assess acceptability and implementation feasibility of the nutrition and PS interventions to be integrated with SAM management. In the present paper, we report findings of qualitative formative research we conducted to develop the content and delivery mode of the PS intervention.

The findings of this formative research informed the development of a PS program that was integrated with SAM management in a pilot study at BMC. The results on impact on child development outcomes and a process evaluation of the contextual factors influencing intervention delivery and uptake will be reported separately.

The study was conducted at the National Institute for Medical Research (NIMR), the Bugando Medical Centre (BMC) tertiary care referral hospital and the Tanzanian Home Economics Association (TAHEA) in Mwanza, Tanzania. Mwanza is located on the shore of Lake Victoria with a population of about 1,182,000 in 2021 (Population Stat, 2022). Main economic activities in the region are agriculture, livestock keeping and fishing. The prevalence of stunting in Mwanza region is 39%, just above the country average of 34%. Among women in the region, 46% have completed primary school, 23% attended or completed secondary school or higher education, and female literacy rates are approximately 77% (Ministry of Health *et al*., 2016). The nutrition unit at the BMC paediatric department manages cases of SAM with complications. The unit provides specialized care and feeding by nurses and a paediatrician. Staff also includes a nutritionist, a social worker and a physiotherapist. Conditions such as sickle cell disease, cerebral palsy and congenital heart diseases are common among malnourished children (Ahmed *et al*., 2016). Outpatient follow-up clinics for malnutrition cases are supervised by a paediatrician and a nurse.

### Participant selection and data collection

Data were collected using in-depth interviews (Guest *et al*., 2013). Caregivers of children, who had recently been admitted with SAM and were receiving outpatient follow-up, were selected by purposive sampling (Campbell *et al*., 2020). Sampling was done with assistance from healthcare staff at the nutrition unit. Caregivers were contacted via phone and invited to BMC or NIMR for interviews. Additionally, in-depth interviews were conducted with professionals from the BMC nutrition unit or NGOs working to promote child development in the community. The interviews with professionals were conducted at their workplaces or community centres. Data triangulation was achieved by interviewing both caregivers of children treated for SAM and professionals working with SAM and ECD.

All interviews were conducted between June-September 2020 and took place in quiet, private environments. They were conducted in Kiswahili by three trained, experienced research scientists from NIMR and lasted 45-60 minutes. Interviews explored awareness and attitudes towards ECD and PS. The interview guides focused on contextual factors that may be barriers or facilitators to supporting the learning and development of a child, as well as current support for ECD in Mwanza. Finally, we gained specific views and suggestions on how a short, focused PS intervention could be designed to improve cognitive development of children during treatment for SAM. Interview guides were pretested and revised to expand probes for deeper exploration as needed. The number of interviews was based on an assessment of data saturation, when no new ideas or themes emerged in interviews (Fusch & Ness, 2015; Saunders *et al*., 2018).

After preliminary data analysis, respondents were invited to a feedback session involving further discussions through which they were able to clarify or elaborate on views and suggestions from the interviews. One group discussion was conducted for the caregivers and another for the professionals. The sessions took 60-90 minutes and were both hosted at NIMR. These sessions were not intended to add new data for analysis but aimed to validate data obtained from the in-depth interviews to ensure correct understanding and to feed-back study findings to participants (Ramanadhan *et al*., 2021).

### Data management and analysis

The interviews were digitally recorded, transcribed verbatim in Kiswahili and translated into English by research assistants at NIMR fluent in both Kiswahili and English. Transcriptions and translations were cross-checked and verified by two authors (ES and GP), fluent in both languages. Transcripts were coded following principles of thematic content analysis (Gale *et al*., 2013) using Dedoose software (version 8.3.35, SocioCultural Research Consultants, Los Angeles, US). First, an initial analytical coding framework was developed deductively by the research team based on previous literature, assumptions and the interview guide. All codes were structured into a codebook with descriptions for each. Then, the first transcript was coded independently by each of the three coders (ES, CJ, HK) and then cross-checked and reviewed by the team to ensure that a similar conceptualisation of framework and codes was used by all coders. Once this preliminary coding was completed, the primary coding framework was created. New codes were created inductively and added to the framework during the following analyses and data familiarisation. Multiple codes were applied to a section of text when appropriate. The remaining transcripts were coded by the coding team and verified by JW, MG and MFO.

Identified barriers and facilitators which emerged from the thematic content analysis were organised and synthesised using the NCF. This framework was selected for its consideration of the different components of care that support caregivers and young children with their development. The barriers and facilitators were examined and linked with one or more of the five NCF components: good health, adequate nutrition, responsive caregiving, opportunities for early learning, and security and safety (WHO *et al*., 2018). Based on this analysis, we report suggestions and opportunities for a PS intervention to improve nurturing care and ECD for children recovering from SAM.

### Ethical Considerations

The study received ethical clearance from the Medical Research Coordinating Committee of the National Institute of Medical Research - Dar es Salaam, Tanzania (Ref-NIMR/HQ/R.8c/Vol.I/1708), and from the ethics committee of the London School of Hygiene and Tropical Medicine (Ref: 17831-1). Written informed consent was obtained prior to starting interviews with all caregivers and professionals. All quotes are used anonymously and without descriptive information that could lead to identification of respondents.

## Results

### Interviewee characteristics

A total of sixteen interviews were conducted. Seven included caregivers (all mothers) of children recently treated for SAM. The majority of caregivers had completed primary or secondary school and were working as small-scale farmers or vendors. The children were between 6 months and 5 years of age, and about half were girls. Most families lived within 10 km from BMC. Nine interviews were conducted with local professionals working at the BMC nutrition unit or at ECD-related NGOs. Professionals included health care workers, nutritionists, and NGO staff. All professionals had more than five years of work experience.

### Barriers and facilitators for providing PS support

#### Good health: the fundamentals of child development

Caregivers and professionals described how illness negatively impacts on children’s growth and ability to learn and should therefore be addressed as a primary focus in any programme to support child development. One mother described how she could see the impact of illness on her child’s development, with similar messages emerging from professionals.

> *“Before falling into sickness, she was so active. She was able to walk around holding objects like tables, chairs. But after falling into sickness, everything stopped* (…) *sickness may slow down the normal growth of a child”* (Mother 7).
>
> *“If a child is frequently under infections* (…) *he will be late to walk, communicate and talk”* (Professional 2).

The health of caregivers is also vital. One professional highlighted how keeping the caregivers’ wellbeing in mind is fundamental as they often have other concerns which make it difficult to take in new messages.

> *“We forget the caregiver’s wellbeing*, (…) *TAHEA comes - they want to train in nutrition or child safety and so on*, (…) *sometimes they* (the caregivers) *don’t listen because they have other problems in mind”* (Professional 6).

Professionals described how increased support of children’s health after discharge from the nutrition unit would be beneficial, in particular by strengthening existing links between the nutrition unit and the community workers.

> *“Community health workers should be trained and become knowledgeable, because us from here* (BMC) *we cannot go there always, we need them, they should be assisting in follow-up”* (Professional 2).

#### Ensuring adequate nutrition and resources at home

Inadequate nutrition was clearly perceived by caregivers as a vital barrier for learning and development of their children.

> *“If the baby doesn’t eat well - probably he cannot grow well, most of these children they are not active* (…) *because even for an adult who doesn’t eat, he or she cannot be active - what about a little child* (…) *eating well is important for child development”* (Mother 7).

Professionals also mentioned adequate nutrition when asked about the foundation for growth and development.

> *“Good nutrition, because he can have all good environment and he is well stimulated, but that won’t be enough if he doesn’t have services like nutrition”* (Professional 5).

Poverty was clearly perceived by both caregivers and professionals as a main barrier for the availability of nutritious food – and as such, a fundamental barrier that cannot easily be addressed by counselling or behaviour change interventions. Mothers demonstrated good awareness of the importance of nutrition and thus experienced the agony of knowing how to feed their child, but not having the resources to do so.

> *“You can think of buying the required food for the child, but you find yourself with no money to buy* (…) *you love your children and wish to buy them everything good for their development, but due to the poor economic status you have no choice”* (Mother 1).

Professional described the poor socio-economic and educational status of the families of children with SAM and emphasized the importance of cooking demonstrations on how to prepare nutritious low-cost meals with locally available foods.

> *“It’s like a mind-set where one knows that to eat well it has to be expensive*, (…) *we are telling them to bring foods that can be found within their environment and we use those foods to cook a meal that contain five groups of nutrients*, (…) *so after you have cooked and prepared a balanced meal, they will say ooh so it’s possible”* (Professional 7).

However, one mother highlighted how the time demand for her income-generating activities resulted in little time for her to keep an eye on her child’s diet, even to a point that led her child to become malnourished.

> *“I was busy with work, actually that is what costed me, because the child was most of the time with the house girl, I had no time to check her diet, then she fell sick, then I was told it was malnutrition. I had to stop my work but it was too late”* (Mother 5).

#### Enabling responsive caregiving through the wider circle of care

Professionals consistently described the importance of having multiple caregivers to support the child and family. This was seen as pivotal for good child-caregiver interactions.

> *“When you find a family which is extended - I mean maybe if there is father, uncle, and others, there is grandmother, those children are so different and their learning and development is very good because they have people to interact and play with”* (Professional 4).

Some professionals highlighted how other caregivers (siblings or housekeepers), who take care of children due to the competing demands on the mother, may be lacking in their ability to provide responsive caregiving.

> *“The mother is busy, a child is raised with a house girl. If the house girl is quiet, a child will learn to be quiet, if she does not get the opportunity to play with other people who can stimulate her”* (Professional 2).

Gender inequities and expectations of gender roles was raised by interviewees as a barrier for responsive caregiving. It was highlighted that society expects mothers to take care of the children whereas fathers are expected to bring money for the household. It emerged from our interviews, that these societal norms may lead to lack of involvement of fathers as they are expected to have other responsibilities and spend little time caregiving.

> *“A father goes to work at 6 am and return home at 11 at night, what time will he play with his child? A child has already gone to bed, he doesn’t have time”* (Mother 2).

Some professionals highlighted their desire to involve fathers and suggested ways to reach fathers through community programmes.

> *“When you go home you will find mothers alone and fathers are not around. So we thought, how can we access fathers* (…). *They are counselled where they are working or using their time* (…), *our counsellors are reporting that they are getting support from men, they are allowed for like twenty minutes while people are taking their coffee while listening to talks about care for child development”* (Professional 5).

Other professionals primarily blamed lack of awareness for insufficient responsive caregiving.

> *“When it comes to playing with a child and understanding a child’s feelings, I don’t think social economics plays a role, I think it’s only knowledge* (…) *To smile for a child doesn’t need money”* (Professional 7).

#### Awareness and prioritisation of play and opportunities for early learning

Professionals and caregivers that we interviewed highlighted how crucial it is to provide young children opportunities to interact and stimulate their senses.

> *“I teach him different things. I play with him, when I get time, I really try my best, even though household chores are all on me”* (Mother 4).

However, professionals also expressed how a lack of awareness of the importance of play leads to lack of prioritisation. A professional described how some caregivers might prioritise cleanliness over social stimulation. Similarly, a mother highlighted lack of understanding as a barrier to PS.

> *“Some kids, their parents stop them from playing with other children, this child then stops playing with others. Parents might think that if the child plays with others, they become dirty not knowing their children’s cognitive development remains stunted as children learn mostly through games”* (Professional 8).
>
> *“I think education matters. Some mothers have not gone to school, she just gave birth to a child - she doesn’t know about playing with a child, maybe she thinks it’s waste of time. I think education should be expanded even in our community”* (Mother 3).

Clearly, time constraints and competing responsibilities may act as barriers towards stimulating the child through play and interaction.

> *“There are some mothers who are vendors, they leave home at 6 am with bucket of tomatoes. She spends a whole day at the market selling tomatoes, when will she play with a child? I have time because I’m a housewife”* (Mother 6).

Professionals suggested ways to promote PS, even in busy families, by integrating it into everyday activities. They gave examples on how to be creative in making toys and stimulate children without need for money.

> *“When the mother performs her duties, does not leave the child, and when she is doing laundry, she tells her child that ‘my child, this is how we do laundry’, so the child knows that when I do this, it is called washing”* (Professional 9).
>
> *“If it is a ball, we look for rags or old clothes, make a ball and tie it properly with rope* (…). *If it is things that make sounds, we will pick gravel, rocks, they will use the rocks to hit on the soda caps, it makes sounds and a child can play with it”* (Professional 8).

#### Issues of security and safety

A young child’s development is vulnerable to physical and psychological stress. In our interviews, although there was limited discussion on this topic, one professional exemplified that stress can impair development and precautions should be taken to ensure the safety of the child.

> *“Another dangerous factor is what we call violent homes, here I mean where at home there is no peace, the parents are fighting and quarrelling and even other relatives, it’s very dangerous for the child’s development”* (Professional 5).
>
> *“You have to observe his safety. There should be playing materials, so you have to ensure there shouldn’t be dangerous objects around. You make sure that environment is safe for a child to learn by himself playing”* (Professional 5).

#### In summary

The factors identified in our interviews include issues that can be addressed by taking a holistic view to creating a PS programme for children treated for SAM, while other factors are more structural in nature and require broader societal initiatives. We have summarised the findings of barriers, suggestions and recommendations identified through our interviews with caregivers and professionals and connected these to the NCF components (Figure 1).

**Figure 1:**
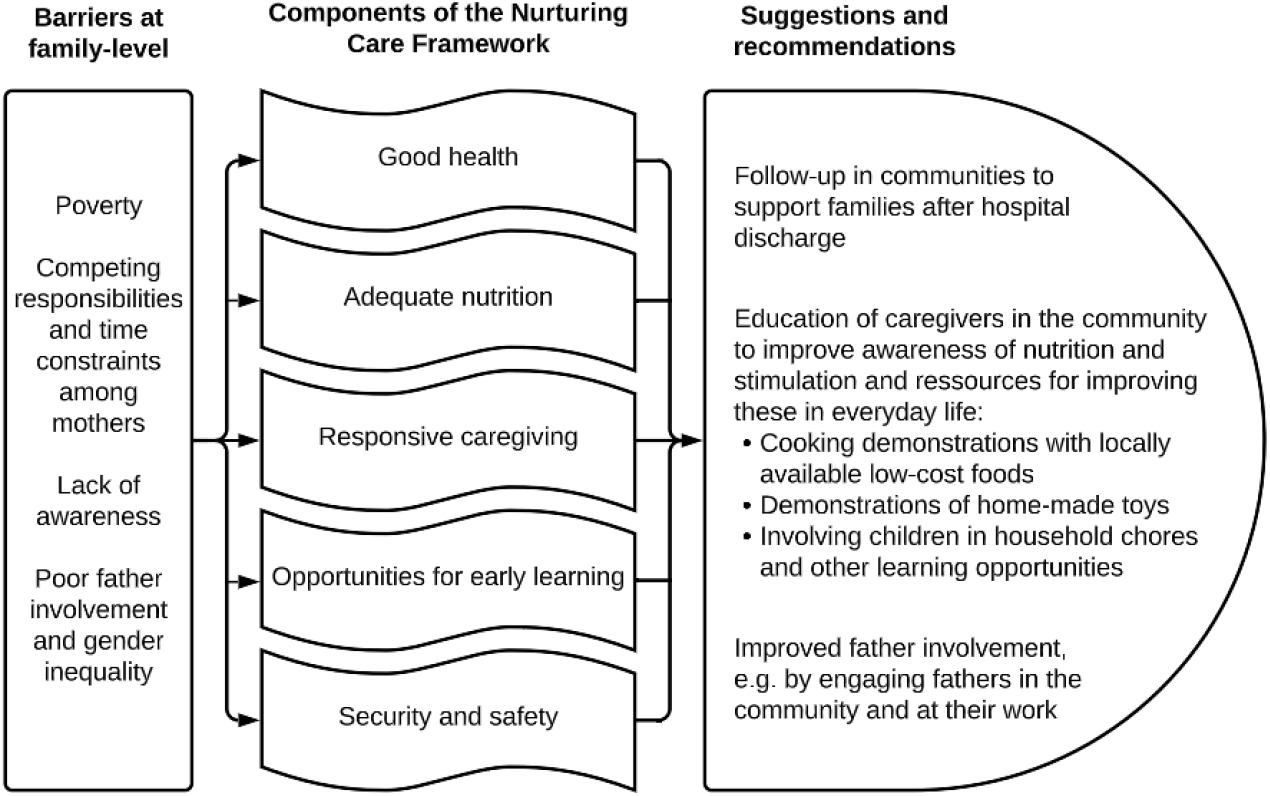
Overview of identified barriers within the family considered to affect nurturing care, and suggestions and recommendations on how to support nurturing care and subsequent learning and development.

## Discussion

This study explored factors to be considered when creating a relevant PS programme for children treated for SAM in a in Tanzanian context. Our results demonstrate social, environment and familial factors that caregivers and professionals believe may influence good nurturing care and which need to be reflected in a programme that could be helpful to families. The findings highlight a number of issues which may inhibit adoption of the five components of the NCF if not considered. These include financial and food insecurity, competing time constraints of caregivers, lack of awareness of the importance of responsive caregiving, play and stimulating activities, and finally, issues of security and safety of children.

Our interviews emphasise how caregivers and professionals are already cognisant of the two NCF components: good health and nutrition, required to meet a child’s developmental potential, but barriers within society or their economy make it difficult for families to support children (Ijaz & Rubino, 2012). Professionals also emphasised the importance of caregiver wellbeing to enable them to provide supportive care for their children (WHO *et al*., 2018). On the other hand, caregivers we interviewed focussed less on the other NCF components: responsive caregiving and opportunities for early learning, which seemed to be less familiar ways of improving children’s development. Another component that was highlighted less in our interviews, was the importance of security and safety. This may need emphasising in counselling on PS support for improving children’s developmental outcomes - however we did not probe specifically into this area and how best to emphasise this might need more consideration.

In this study, we found that poverty and financial insecurity were commonly identified as barriers to provide key components of nurturing care; including adequate nutrition and opportunities for early learning. Mothers are generally knowledgeable on what constitutes good nutritional support for children, but many families have limited resources to act on this knowledge. Some professionals interviewed suggested that cooking demonstrations might improve understanding of how to provide better nutrition at a low cost from available ingredients. Professionals also suggested that educating mothers in low- and no-cost strategies for creating opportunities for early learning, would be possible. This includes encouraging families to make home-made toys from available materials and engaging children in household activities as a form of play. These suggestions were found relevant to our context in Tanzania and are likely to be generalizable broadly across low-income settings (Gelli *et al*., 2018; Grantham-McGregor *et al*., 1987). However, the evident impact of poverty, particularly among families who bring their children into a nutrition unit, underlines a need for additional support through cash transfer programmes, social welfare systems, or similar – some of which have demonstrated evidence of improving child development (Attanasio *et al*., 2014; Evans *et al*., 2019). It is widely documented that poverty has a multi-faceted and detrimental effect on child development (Fernald & Gunnar, 2009; Fernald *et al*., 2011) and we acknowledge the fundamental need for these broader issues to be addressed through structural changes, although these lie beyond the scope of our present programme.

Gender norms in the study setting may mean that women are responsible for household duties including raising children and men expected to be key earners (Feinstein *et al*., 2010). Our interviewees projected responsibility for responsive caregiving on mothers and rarely mentioned other people within the circle of care. Many mothers who we interviewed described feelings of being overwhelmed and lacking time for additional activities. This may be common across the globe. Analysis of the UNICEF multiple indicator cluster surveys found that, across 38 low-income countries, 48% of fathers did not engage in any stimulation activities with their child (Jeong *et al*., 2016). Furthermore, a recent meta-analysis of 102 interventions to improve ECD outcomes found only 7% involved fathers in the intervention and only one study assessed paternal stimulation outcomes directly (Jeong *et al*., 2021). To shift this, it is likely that programmes would need to address cultural expectations of a father’s role as well as the wider community’s (including men’s) understanding of the importance of responsive caregiving for children’s brain development. It is our recommendation that this be considered in future programmes within community settings.

Many mothers of children suffering from acute malnutrition in the Mwanza region have no or low level of education (Ahmed *et al*., 2016). Studies have demonstrated that maternal schooling is strongly associated with nutritional outcomes of their children and that maternal knowledge regarding causes of malnutrition is a strong predictive factor of childhood stunting (Ahmed *et al*., 2016; Hasan *et al*., 2016). It is widely reported that maternal schooling has a strong positive effect on child development (Cuartas, 2021). Whilst the majority of these studies are from high income settings, a recent study in Uganda found that for each additional year of maternal education, the child’s ECD index score rose by 0.12 increments, which translated into effects on ability to read, identify letters and numbers at age 3-4 years (Cuartas, 2021). This suggests that not only educational interventions to improve maternal knowledge of PS and nutrition could prove effective for children suffering from SAM in rural Tanzania, but more so, supporting girls to remain in school, might also be fundamental. Clearly approaches to improve ECD should holistically encompass the needs of families, healthcare staff and communities and consider the wider structural and social determinants of health. This is complex and will require working with ministries and agencies who may be able to support this on a wider scale. Although these aspects are not addressed further within our specific programme in the nutrition unit, we hope that by continuing to highlight themt, we can advocate for programmes which start early with support for young girls and women to stay in education.

Suggestions from caregivers and professionals on how to overcome some of the barriers highlighted included recommendations that could be implemented into a short, focussed intervention during management of SAM. These included better linkage with community workers on discharge from the nutrition unit and continued education of caregivers to improve knowledge of both nutrition and PS. Professionals provided specific recommendations on ways to provide advice to caregivers which could be used in everyday scenarios, e.g. cooking demonstrations, homemade toys and encouraging learning activities when involving children in everyday chores. Whenever possible, getting fathers more involved in care as well as generally widening the circle of care may make a big difference to enabling children to grow and develop better.

Earlier studies suggest various intervention strategies and programme models, which can be used to deliver parenting interventions and achieve improved ECD (Jeong *et al*., 2021). In particular, interventions provided early and more intensively (e.g. weekly) with supportive supervision may be most effective (Aboud *et al*., 2018). The majority (70%) of published effective parenting interventions last at least 12 months (Jeong *et al*., 2021) although several meta-analyses have shown no significant correlations between programme duration and ECD outcomes (Jeong *et al*., 2021; Sanders *et al*., 2014) or between more parent contacts and larger effect sizes (Engle *et al*., 2011).

### Strengths and limitations

Our findings derive directly from caregivers and professionals who work with ECD or whose children have SAM in Mwanza, Tanzania. These may be of relevance for other low-income settings, especially in sub-Saharan African countries. We are aware that the issues raised may be subject to some degree of social desirability bias, and the reported practices and perceptions may not necessarily correspond to actual behaviours. We hope that this study may shed light on some issues that could be addressed in creating stronger support for children to thrive if provided with both nutritional and PS support in a programme embedded with a clear understanding of the wider social and structural issues which affect the care of a child admitted to the nutrition unit. Our qualitative approach has allowed us to gain a nuanced perspective on barriers for learning and development in young children by gaining points of view from both caregivers and professionals which we can share and learn from. Additionally, inclusion of professionals working at different levels within the hospital and at different NGOs contributed to a broader variety of perspectives.

## Conclusions

Based on the barriers, facilitators and suggestions we identified, we found that a context-relevant PS programme to improve child development among young children treated for SAM in Tanzania should include better linkage with community workers on discharge from the nutrition unit, education of caregivers to improve awareness of nutrition and PS, and involvement of fathers to widen the circle of care and decrease the burden on mothers. Specific recommendations from professionals on counselling of caregivers which could be useful in everyday scenarios included cooking demonstrations, homemade toys and encouraging learning activities by involving children in everyday chores. Other issues identified were structural, highlighting the continued need for other types of programmes to target food and financial insecurity.

## Data Availability

Data available on request due to privacy/ethical restrictions

## Acknowledgements

The authors would like to thank the interviewees who took time to participate in this study and share their views and thoughts with us. We would also like to acknowledge the healthcare staff at the Bugando Medical Centre nutrition unit, who helped sampling and identifying the caregivers for interviews, as well as the staff at the National Institute for Medical Research, who also participated in this study.

